# Suicide and mental health during the COVID-19 pandemic in Japan

**DOI:** 10.1101/2020.10.06.20207530

**Authors:** Michiko Ueda, Robert Nordström, Tetsuya Matsubayashi

## Abstract

**Background:** The coronavirus disease (COVID-19) pandemic is an unprecedented public health crisis, but its effect on suicide deaths is little understood.

**Methods:** We analyzed data from monthly suicide statistics between January 2017 and October 2020 and from online surveys on mental health filled out by the general population in Japan.

**Results:** Compared to the last three years (2017-2019), the number of suicide deaths was lower than average during the initial phase of the pandemic but exceeded the past trend starting in July 2020. The source of the increase was mainly female suicides whose numbers increased by approximately 70% in October 2020 (IRR: 1.695, 95% CI: 1.558-1.843). The largest increase was found among young women (less than 40 years of age). Our survey data indicated that the status of mental health among young women has been deteriorating in recent months, and that young female workers were more likely than any other group to have experienced a job or income loss, suggesting adverse economic conditions surrounding some of these individuals.

**Conclusions:** Our results indicate that continuous monitoring of mental health, particularly that of the most vulnerable populations identified in this study, and appropriate suicide prevention efforts are necessary during the COVID-19 pandemic.

## INTRODUCTION

The coronavirus disease (COVID-19) pandemic is an unprecedented public health crisis that has both physical and mental health consequences. Emerging evidence suggests that the COVID-19 pandemic severely affects the mental health of the general population.^1,2^ These findings correspond with concerns that deteriorating mental health, in combination with higher unemployment rates during the pandemic, have the potential to increase the incidence of suicide worldwide.^3,4^

Despite this risk, to the best of our knowledge, no peer-reviewed study has documented how the COVID-19 pandemic has affected the prevalence of suicide at the national level by major demographic groups, owing to the lack of reliable fast-reporting data. The present study examined the suicide deaths and mental health status of the general population during the COVID-19 pandemic in Japan. We focused on Japan for two reasons. First, Japan has one of the highest suicide rates among OECD countries.^5^ Its suicide rate (the number of suicide deaths per 100,000 people) in 2018 was 16.5 and in that year, more than 20,000 people ended their lives by suicide.^6^ Thus, the vulnerable population in Japan may be highly responsive to the pandemic-inducted crisis, which could result in a rapid increase in suicide deaths. Second, Japan has rapid-release monthly suicide statistics, which, in combination with our own monthly survey data, allow us to closely monitor the status of the general population since the onset of the pandemic.

Japan had its first encounter with COVID-19 earlier than most of the rest of the world. The first case was reported on January 16, 2020, followed by outbreaks on a cruise ship at Yokohama Port in February 2020. The first wave of COVID-19 cases was observed in April 2020, and a second, larger wave of cases started in July 2020. As of October 31, 2020, the total number of confirmed COVID-19 cases was 91,329, and the number of deaths attributed to COVID-19 was 1,754, or 13.95 deaths per 1 million people.^7^ The number of new COVID-19 cases in Japan is shown in Supplementary Figure 1.

The Japanese government has imposed several measures to stem the tide of the pandemic. School closure started in early March. The Japanese government declared a state of emergency in major metropolitan areas on April 7, 2020, which was extended to the rest of the country on April 16. Without introducing lockdown measures or strict domestic movement restrictions during the state of emergency, authorities requested non-essential businesses to close or opt to work remotely. Stores and restaurants were asked to operate for reduced hours. The state of emergency was lifted on May 25, 2020.

## METHODS

We analyzed the publicly available monthly suicide statistics tabulated by the National Police Agency (NPA) and published by the NPA and the Ministry of Health, Labour and Welfare of the Japanese government.^8,9^ The NPA suicide statistics are based on police investigations of suicide cases. In Japan, the police investigate all suspected cases of suicide, and the agency records all deaths that are determined to be suicide in its suicide statistics data. We used the agency’s monthly tabulation between January 2017 and the most recent available data, October 2020 (as of November 30, 2020). There are two types of NPA data: those tabulated based on the date that the deceased was found and those tabulated by the date of death. We used the former, as these is the most frequently updated data.

We examined the total number of deaths by suicide, the number of suicides by sex, age group (less than 40 years, 40–59 years, and 60 years and older), and major occupation. We estimated the following Poisson regression model:

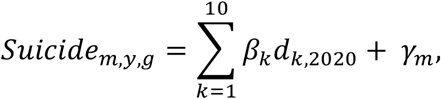

where the dependent variable is the number of suicides in month *m* in year *y* for group *g*, where *m* = 1, …, 12. *d*_*k*,2020_ is an indicator variable for each month in 2020 (until October), where *k* = 1, …, 10 and its associated coefficient, β_*k*_, captures the difference in the number of suicides in month *k* relative to the baseline period, which is the corresponding month in the last 3 years. The month fixed effects (i.e., 12 indicator variables for each month), γ_*m*_, were included to capture the baseline monthly fluctuations in suicide deaths. To facilitate the interpretation of the Poisson regression results, we converted the coefficients to incidence rate ratios (IRR). This part of the analysis used publicly available data; thus, no ethics approval was necessary.

In addition, we assessed the status of mental health among the general Japanese population by using data from a series of monthly online surveys that we have been conducting since April 2020. We selected the respondents from a commercial online panel so that they were representative of the Japanese population in terms of sex, age group, and area of residence. Each monthly survey contained 1,000 respondents. A more detailed description of the survey is provided in Supplementary File 1. For the purposes of this study, we used surveys taken between April and October 2020 (N=7,000).

As measures of mental health, we analyzed the prevalence of depressive and anxiety symptoms, measured by the Patient Health Questionnaire nine-item scale^10^ and the seven-item General Anxiety Disorder scale (GAD-7),^11^ respectively. We also examined the self-reported change in employment status of the respondents in the labor market to understand whether they had experienced major job-related changes during the COVID-19 pandemic. We asked about changes in employment status only since June, and we used the data between June and October 2020, when we analyzed the job-related changes of the respondents (N=5,000). We calculated the presence of depressive symptoms (PHQ-9 ≥ 10) and anxiety symptoms (GAD-7 ≥ 10) for all respondents, as well as the percentages of those in the labor force who had lost their job, taken leave or been laid off from work, or experienced significant reduction in working hours in the last 3 months, and stratified them by sex and age group.

The survey was approved by the Ethics Review Committee on Human Research of Waseda University (approval #: 2020-050) and Osaka School of International Public Policy, Osaka University.

## RESULTS

During our study period, the number of suicides recorded in the NPA data was 81,431, of which 55,963 cases were male and 25,468 suicide cases were female. The average number of suicides per month was 1,732.57 (SD = 158.98), and the maximum number was 2,158 which was recorded in October 2020.

The top panel of Figure 1 shows the IRRs for the months in 2020 when we used the number of suicide deaths in Japan overall and by sex as the outcomes. The estimation always includes indicator variables for each month, and thus, the baseline period is the mean of 2017–2019 in the corresponding months. The number of suicide deaths in the two time periods is provided in Supplementary Figure 2. The shaded area in the figure indicates the period of the state of emergency. Starting in February, when the country had a major encounter with the disease with the Diamond Princess case, Japan experienced a decrease in suicide deaths. However, the declining trend observed in the early phase reversed in July and the IRR increased to 1.302 in October (95% CI: 1.237–1.369). The source of the increased suicide cases was mainly female suicides; the IRR for female suicides was 1.181 (95% CI: 1.079–1.292) in July, and it continued to rise until October (IRR: 1.695, 95% CI: 1.558–1.843).

**Fig. 1:**
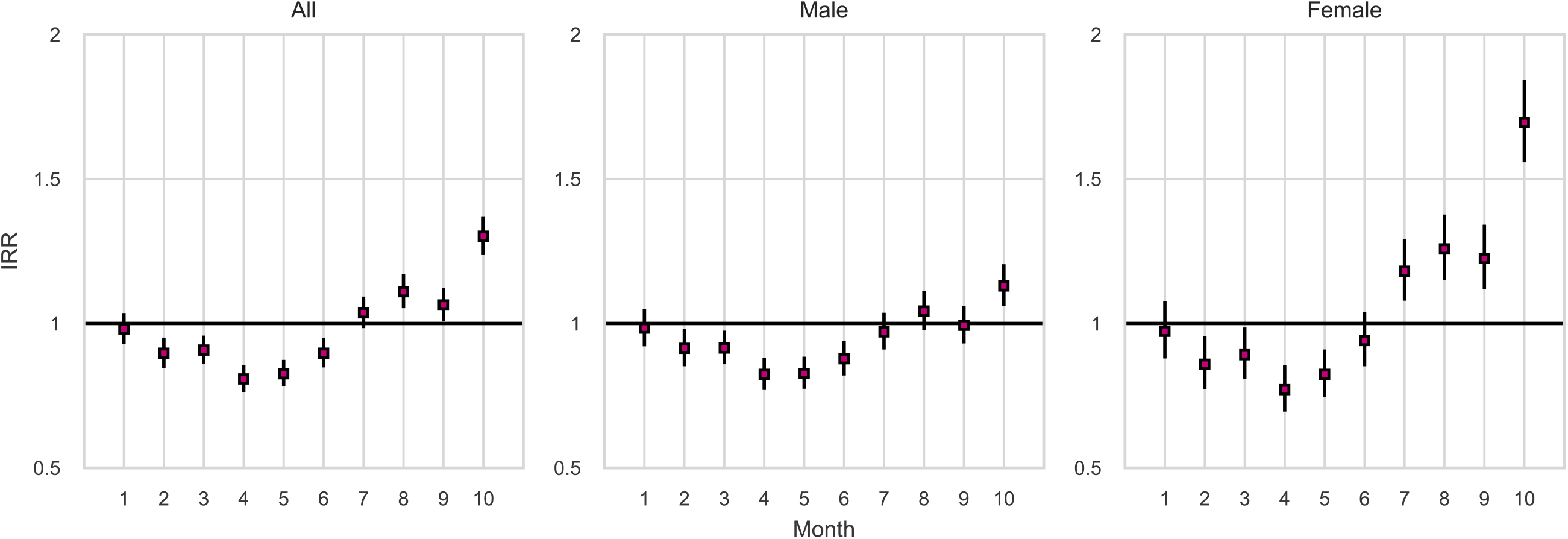
The incidence rate ratios for suicides in Japan in 2020 relative to 2017-2019: total and by sex.

According to Figure 2, which reports the IRRs by sex and age group, suicide deaths by all demographic groups exhibited the same trajectories, with an initial decline during the state of emergency, followed by an increase starting in July. We observed the largest increase among young individuals (less than 40 years old) for both sexes. In particular, the incidence of suicide among young women was 1.648 and 1.956 (95% CI: 1.384–1.962 and 1.66–2.304) for August and October, respectively. However, in October, suicides by middle-aged women (age group 40–59 years) also sharply increased (IRR: 1.955, 95% CI: 1.679–2.276).

**Fig. 2:**
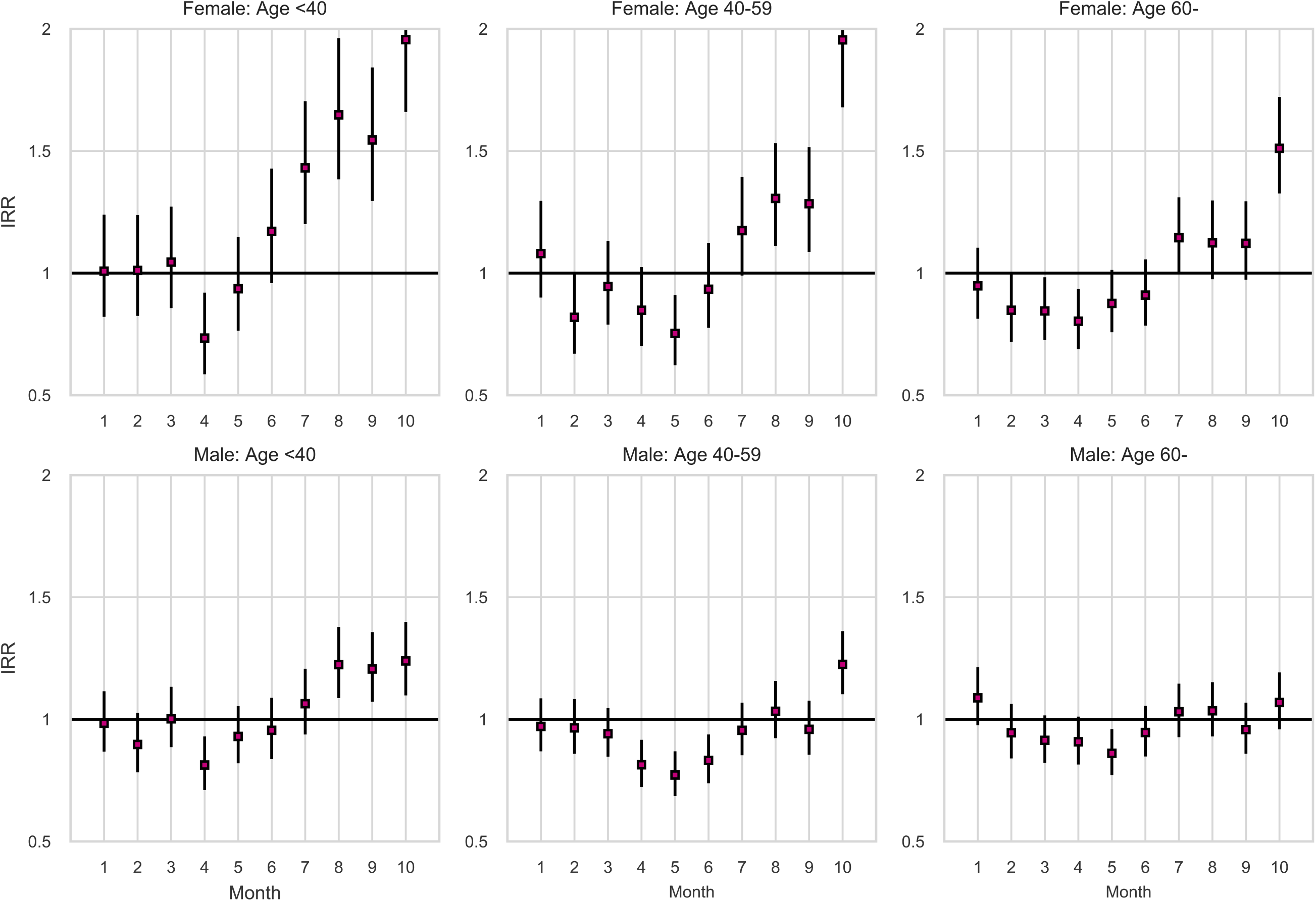
The incidence rate ratios for suicides in Japan in 2020 relative to 2017-2019: by sex and age group.

Figure 3 reports the IRRs for the months in 2020 by major occupation categories. The number of suicide deaths by occupation group is provided in Supplementary Figure 3. It indicates that students and homemakers experienced the largest increase in suicides during the second phase of the pandemic. Student suicides started to increase in June with its IRR peaked in August (1.915, 95% CI: 1.678–2.186). The increase was observed not just among middle and high school students; the number of suicides by university students almost doubled in August and September 2020 (46 and 51 in 2020, respectively, the average in 2017-2019 in each month: 24). The incidence of suicide among homemakers also increased starting in the summer months, with the IRR for homemaker suicides rising to 1.873 in October (95% CI: 1.661–2.113). Among the employed, the number of suicides started to rise in July and the IRR in October was 1.335 (95% CI: 1.271–1.402).

**Fig. 3:**
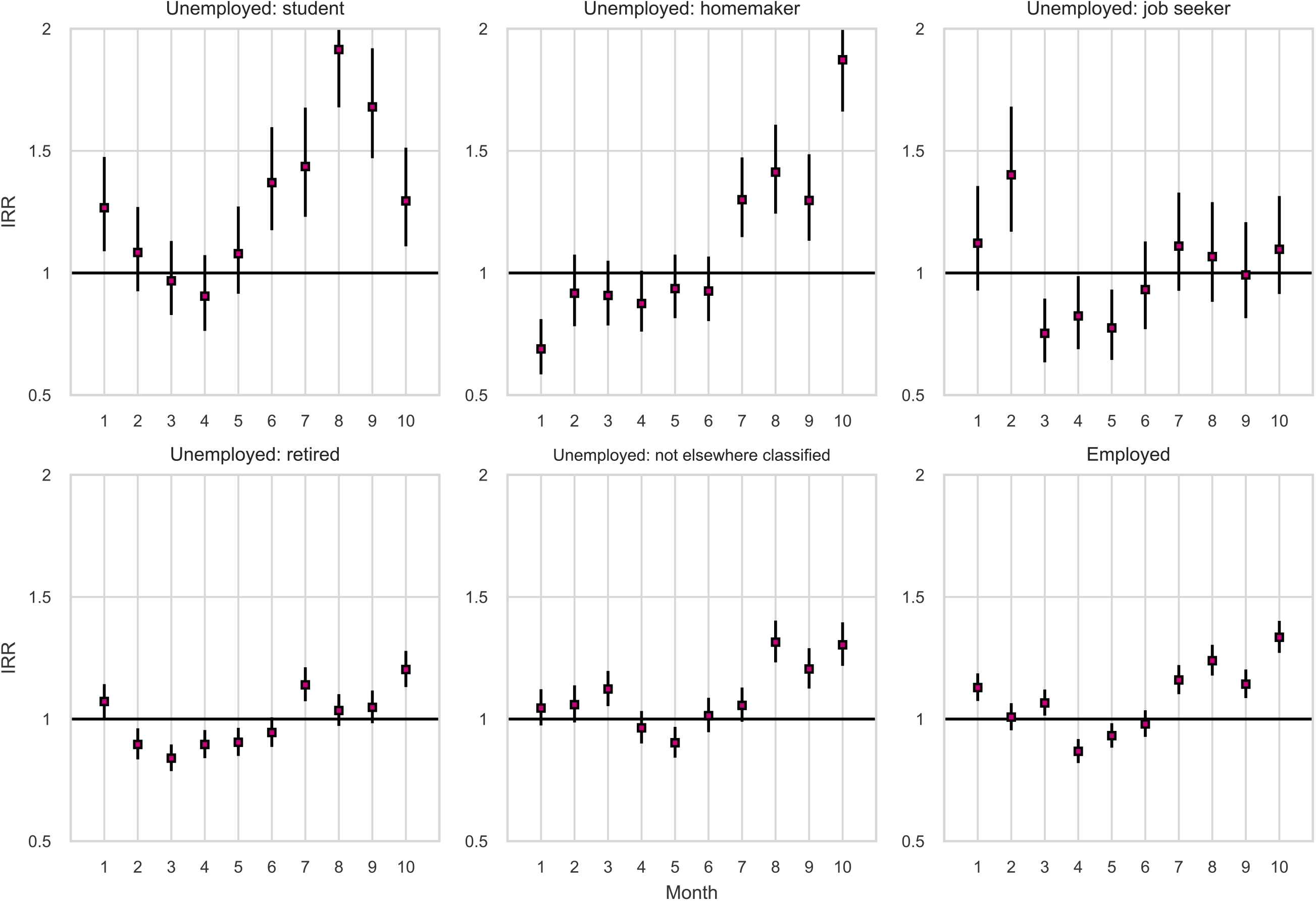
The incidence rate ratios for suicides in Japan in 2020 relative to 2017-2019: by major occupation group.

Figure 4 reports the percentages of those categorized as having depressive symptoms or anxiety disorders according to sex and age group. The top panel shows the prevalence of depression based on PHQ-9 score, and the bottom panel shows the percentages of those who were considered to have anxiety disorders based on their GAD-7 score. Overall, relatively young individuals (those less than 40 years old) were more likely to have depressive and anxiety symptoms than those in older age groups. The June data reported the worst status of mental health for these relatively young individuals, which was right after the state of emergency was lifted. Figure 4 also suggests that the prevalence of depressive symptoms among relatively young women has been increasing since July, with 31.08% of them (N = 148) being classified as depressed in October.

**Fig. 4:**
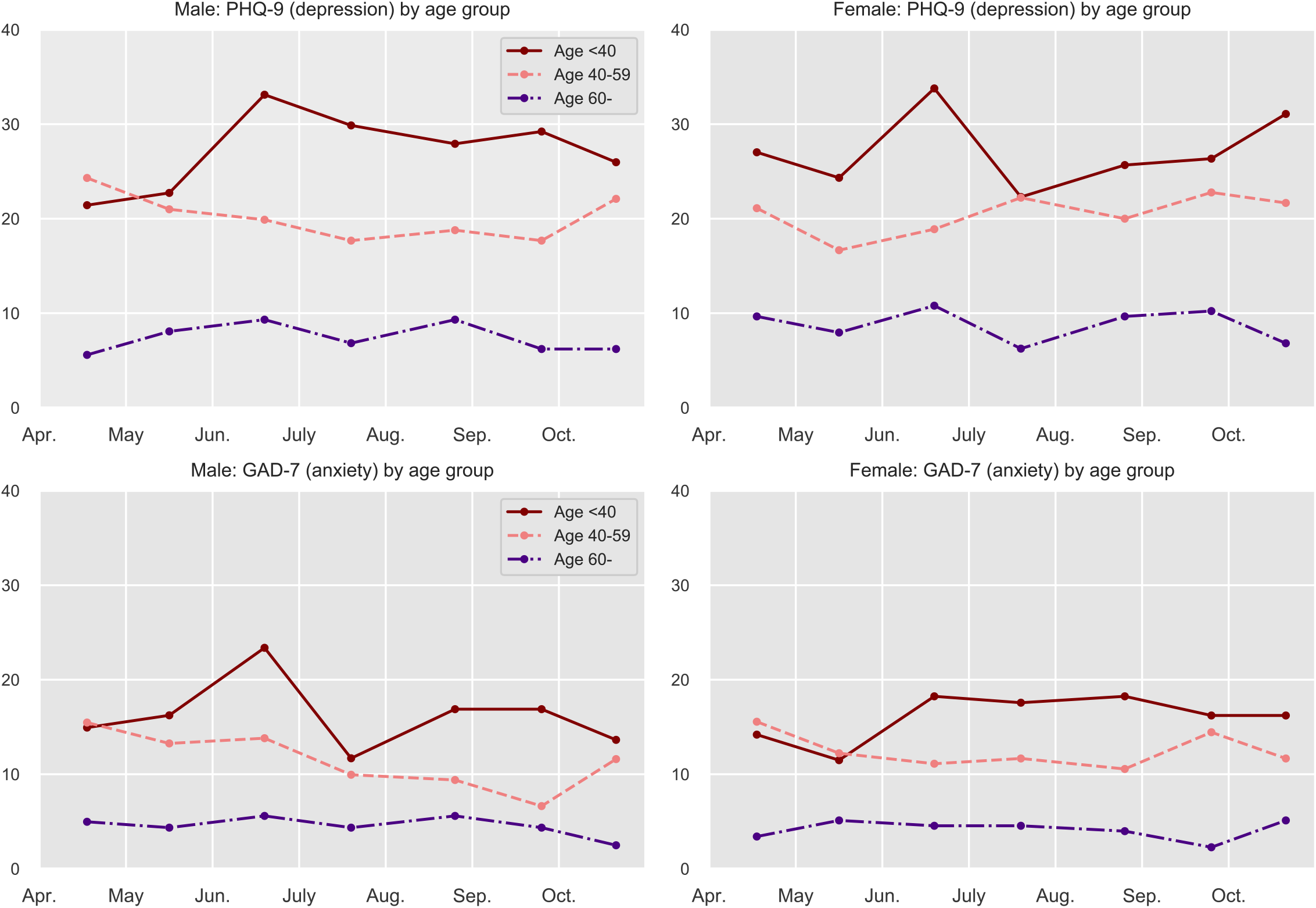
The prevalence of depressive/anxiety symptoms among the general population in Japan by sex/age group: April - Oct. 2020. Note: N=7,000. PHQ-9: the 9-item Patient Health Questionnaire. GAD-7: the 7-item Generalized Anxiety Disorder Scale. PHQ-9 score of 10 or higher or GAD-7 score of 10 or higher was used as a cutoff.

Finally, when we analyzed our survey respondents in the labor force who reported that they had lost their job, taken leave/been laid off from work, or experienced a drastic decrease in working hours over the past 3 months, stratified by sex and age groups, female respondents were more likely to have reported changes in their employment status and working hours, with 21.37% of them reporting such changes, compared to 16.25% of male workers. Among female workers, 25.24% of young individuals (less than 40 years old) reported drastic changes in employment and working conditions, whereas the corresponding number for young male workers was lower at 17.01%. The full data are reported in Supplementary Table 1.

## DISCUSSION

This study examined the monthly trajectories of suicide deaths in Japan during the COVID-19 pandemic. We found that the number of suicide deaths was lower during the initial phase of the pandemic in 2020 than the average during the period of 2017–2019. Starting in July 2020, however, the number exceeded the trend of the past 3 years. Across all age and occupation groups, the largest increase was found among relatively young women, those aged less than 40 years old at the time of their death. The prevalence of suicide among students and homemakers during the second phase of the pandemic was also notably higher in 2020 than the monthly averages of 2017–2019.

Potential factors behind the large increases in suicide cases by women and young individuals during the second phase of the pandemic include the economic consequences of the pandemic, school closure, and media reporting of celebrity suicides. The economic damage of the current pandemic was particularly evident in industries that are served mainly by women, such as the service, retail, and travel industries.^12^ The labor statistics in October 2020 indicate that the number of employed individuals in non-permanent positions, such as part-time or contract workers, decreased for eight consecutive months from the onset of the pandemic. Between July and October 2020, the number of these precarious workers decreased by 4.59 million from the same months in the previous year, of which 2.91 million were women.^12^ It should also be noted that the occupation categories reported in the NPA data are based on the occupation at the time of death, and some women might have been categorized as homemakers if they had lost their jobs prior to their death.

The results of our survey also revealed the unfavorable psychological and economic conditions of young women. We found that the status of mental health among young women (less than 40 years of age) was worse than that of women aged above 40 years, which may underlie their relatively high suicide rate in the second half of 2020. Relatively worse mental health conditions among young and economically vulnerable individuals were also reported in another study, in which other attributes of the respondents were explicitly taken into account.^13^ In addition, we found that young female workers were more likely to have experienced a job or income loss in recent months than any other groups were, suggesting their adverse economic conditions.

The increase in student suicides might be associated with school closure and an unusual school calendar in 2020. Schools in Japan closed around March 2, after the former Prime Minister, Shinzo Abe, abruptly announced a request for school closure on February 27, 2020, and they did not reopen until the state of emergency was lifted in late May. Thus, elementary to high school students had to stay at home for almost 3 months. Emerging evidence indicates the negative impact of school closure on students; a study conducted between June and July (after school reopening) found that 72% of surveyed students (age 7–17 years) reported symptoms that indicated some form of stress reaction.^14^

Returning to school after a long break is known to be difficult for some students even in normal years, and student suicides have previously peaked on the first day of school after the summer break, specifically on September 1.^1^ Elementary to high schools in Japan resumed in around late May and reopened in August after a short summer break in 2020, which may explain the observed increase in student suicides starting in June.

As for university students, most universities in Japan switched to online teaching when the new academic year started in April, and largely stayed online during the spring and fall semesters.^16^ It is possible that some university students were under distress because online and solitary learning without on-campus activities continued for a prolonged period; 29.41% of the students in our survey (N = 153) were classified as depressed, equivalent to the prevalence of depression symptoms among those who had lost their jobs (Supplementary Table 2).

Media reporting on celebrity suicides might also explain some of the increases in suicide deaths observed in this study. Several well-known individuals died by suicide during the pandemic; two notable instances were the suicide death of a male actor, who died at the age of 30 years on July 18, 2020, and that of a 40-year-old actress on September 27, 2020. Both of them were at the peak of their career and the media reporting on their unexpected suicide deaths might have affected vulnerable individuals. The subsequent increase in actual suicides is known to be particularly large when celebrity suicides are widely discussed on social media or when they are regarded as a “surprise,”^17,18^ both of which are applicable in the abovementioned two cases.

Although the number of suicides started increasing in July 2020, Japan experienced a decline in suicide deaths in the initial phase of the pandemic. The decline may be explained by enhanced social connectedness during times of crisis. For instance, the level of social connectedness and altruism is known to increase in the aftermath of natural disasters,^19^ and if natural disasters and the current pandemic share certain similarities, then enhanced connectedness might have worked as a protective factor with regard to suicide risks.

This study has several limitations. First, our analysis used provisional monthly data on suicide deaths, which may be corrected upward in subsequent months. Thus, any reported decline should be interpreted with caution. Second, our data on suicide deaths are aggregate in nature, because the NPA does not release individual-level data; thus, our analysis could not control for the impacts of confounders. Third, our online surveys relied on the commercial panel of respondents, who were not randomly sampled from the general population. Thus, the survey data might not include certain segments of the population.

Despite these limitations, the present study makes an important contribution to the scientific community by reporting the trajectories of suicide deaths during the COVID-19 pandemic and by highlighting the most vulnerable populations during this unprecedented public health crisis. The experience in Japan may provide valuable implications for other countries. Given that the most affected industries were those mainly served by women in many other countries, it is likely that women constitute one of the highest risk groups for suicide in those countries as well. Similarly, since many countries also introduced school closures during the peak period of the pandemic, future studies should investigate the impact of school closures on schoolchildren and university students. The results of our study strongly indicate that continuous monitoring of mental health and appropriate suicide prevention efforts are necessary during and after the COVID-19 pandemic.

## Data Availability

The data on suicide deaths are public data. The individual survey data cannot be shared due to the sensitive nature of the study.

## Supplementary Information: Description of the online surveys

The status of mental health and economic conditions of the general population reported in this study were based on a series of ongoing monthly surveys. We sampled our respondents from a web panel of a major commercial survey company in Japan, the Survey Research Center. For each survey, the company sent out screening questions to approximately 10,000 registered individuals and then constructed a final sample of 1,000 respondents so that they were representative of the Japanese population in terms of their sex, age groups, and areas of residence.

Each of the surveys was conducted on the following dates:

1^st^ survey – April 16-18

2^nd^ survey – May 15-17

3^rd^ survey – June 17-19

4^th^ survey – July 17-21

5^th^ survey – August 24-26

6^th^ survey – September 23-25

7^th^ survey – October 20-22

**Supplementary Table 1:** The reported experience of major change in employment status or working hours by sex and age group: June - October 2020.

|  | **All** | **Experienced major change in employment status or working hours** |  |
| --- | --- | --- | --- |
|  | **N** | **N** | **%** |
| Total | 3813 | 708 | 18.57 |
| Female, Age <40 | 618 | 156 | 25.24 |
| Female, Age 40-59 | 720 | 146 | 20.28 |
| Female, Age 60- | 389 | 67 | 17.22 |
| Female (all ages) | 1727 | 369 | 21.37 |
| Male, Age <40 | 729 | 124 | 17.01 |
| Male, Age 40-59 | 869 | 129 | 14.84 |
| Male, Age 60- | 488 | 86 | 17.62 |
| Male (all ages) | 2086 | 339 | 16.25 |

Note: The data represent respondents from the four original surveys taken between June and October 2020. Only respondents in the labor force (N=3,813) were included in the tabulation. The exact question asked in the survey was “During the past three months, have you experienced a job loss, a layoff/temporary leave of absence, or significant reduction in working hours?”

**Supplementary Table 2.** The prevalence of depressive and anxiety symptoms by employment status.

|  | **N** | **% PHQ9 ≥10**<br>**(depression)** | **% GAD7 ≥10**<br>**(anxiety)** |
| --- | --- | --- | --- |
| Permanent employee | 2599 | 20.47 | 11.35 |
| Part-time/temporary worker | 741 | 21.72 | 13.23 |
| Self-employed | 265 | 13.58 | 5.66 |
| Unemployed, laid off, on leave | 386 | 30.57 | 21.50 |
| Not in the labor force | 2856 | 13.59 | 7.98 |
| Student | 153 | 29.41 | 16.99 |
| All | 7000 | 18.29 | 10.64 |

Note: The data represent respondents from the five original surveys taken between April and October 2020. PHQ-9: the 9-item Patient Health Questionnaire. GAD-7: the 7-item Generalized Anxiety Disorder Scale. PHQ-9 score of 10 or higher or GAD-7 score of 10 or higher was used as a cutoff.

**Supplementary Fig. 1:**
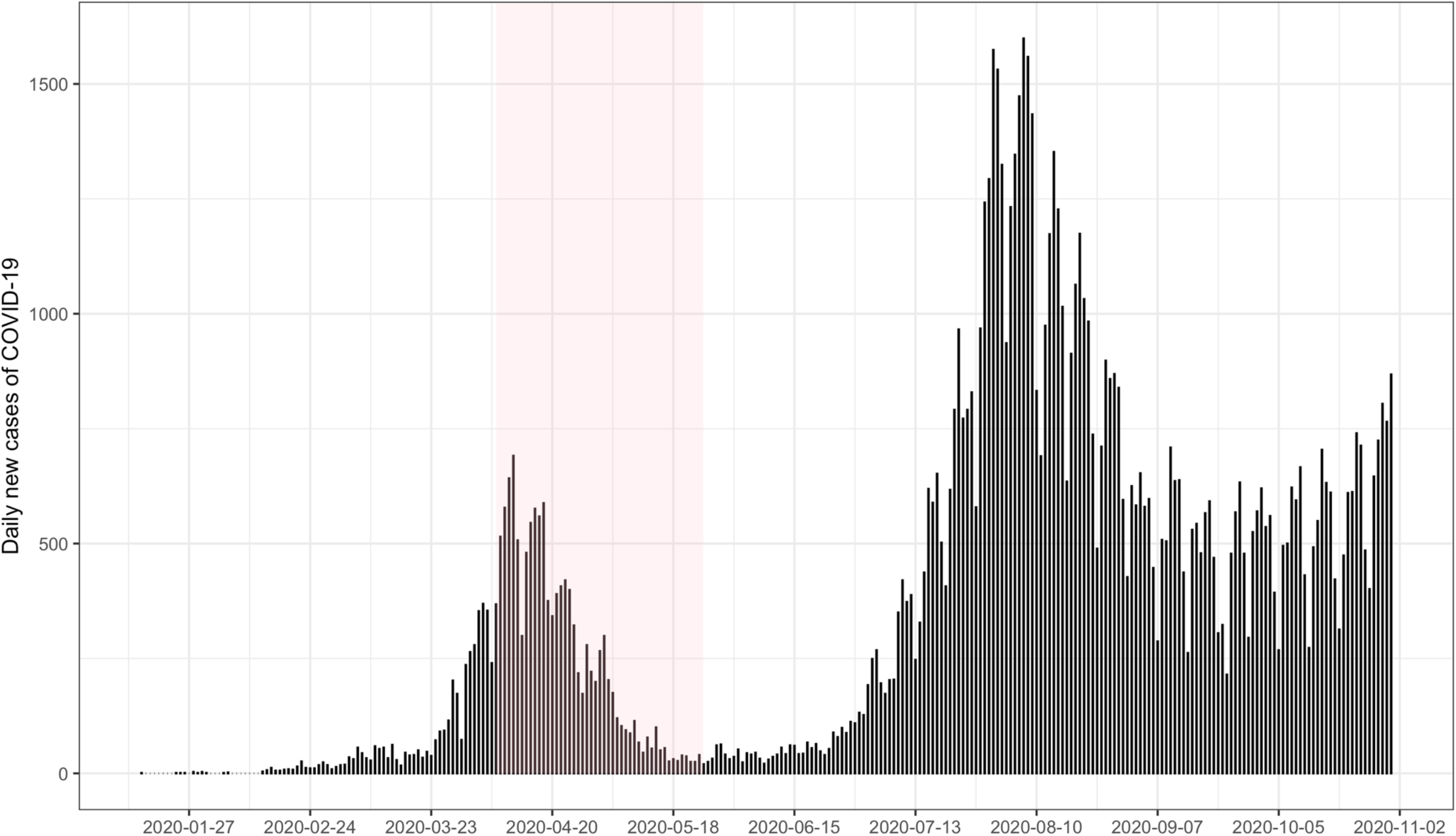
The number of new COVID-19 cases in Japan. Note: The shaded area indicates the period of the state of emergency (April 7-May 25).

**Supplementary Fig. 2:**
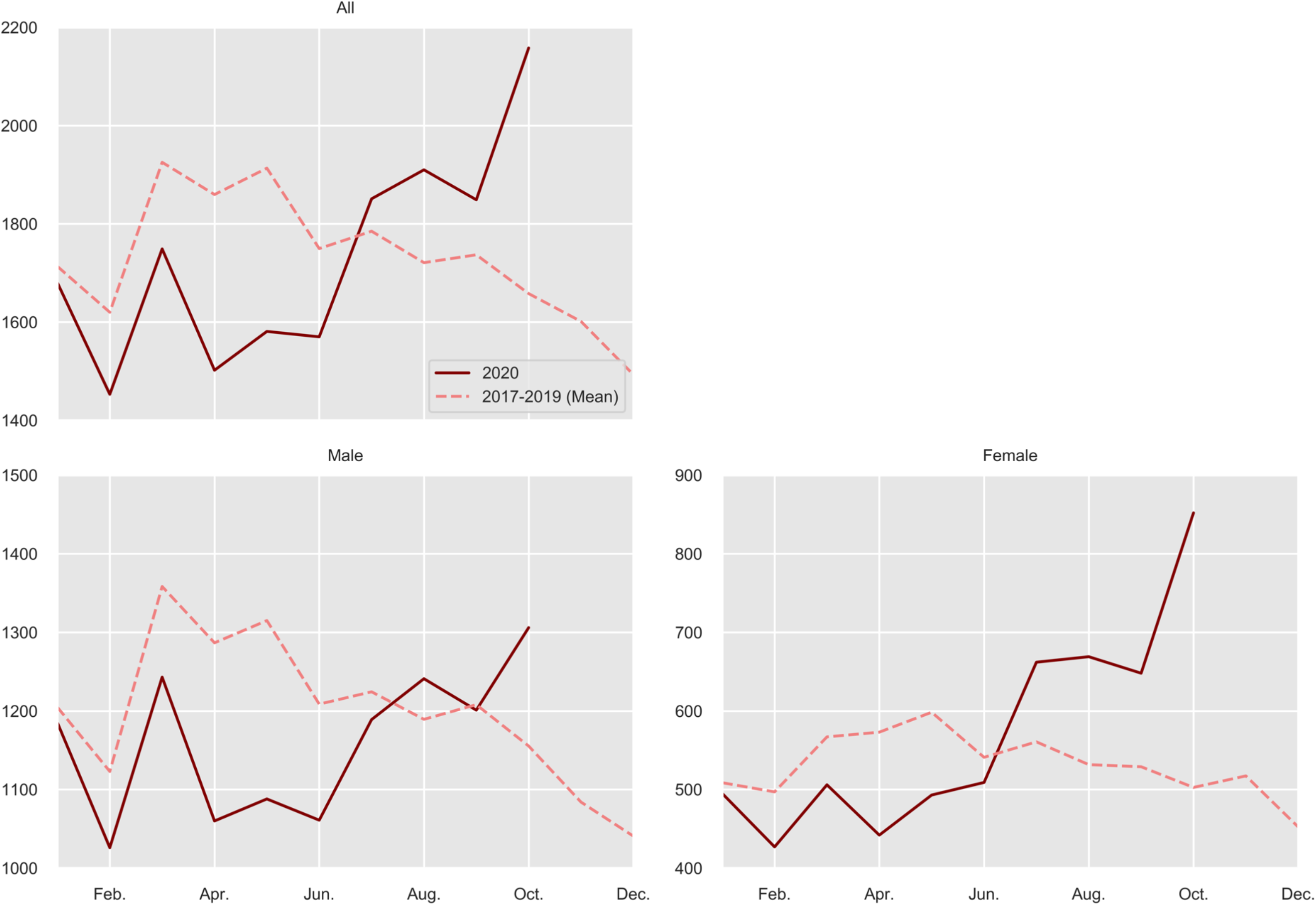
The number of suicides in Japan in 2020 and 2017-2019.

**Supplementary Fig. 3:**
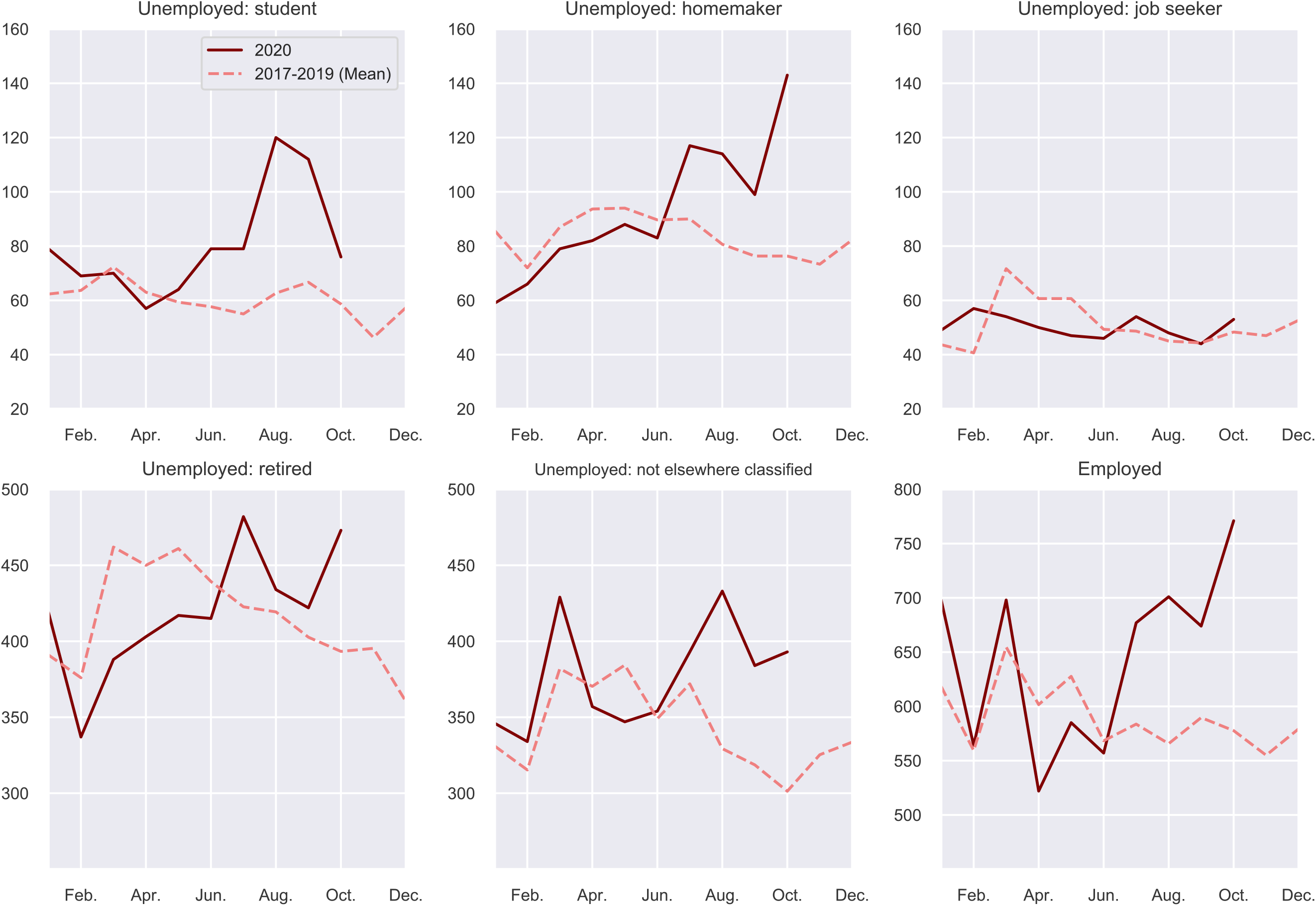
The number of suicides in Japan in 2020 and 2017-2019 by major occupation group.

**Supplementary Fig. 3:**
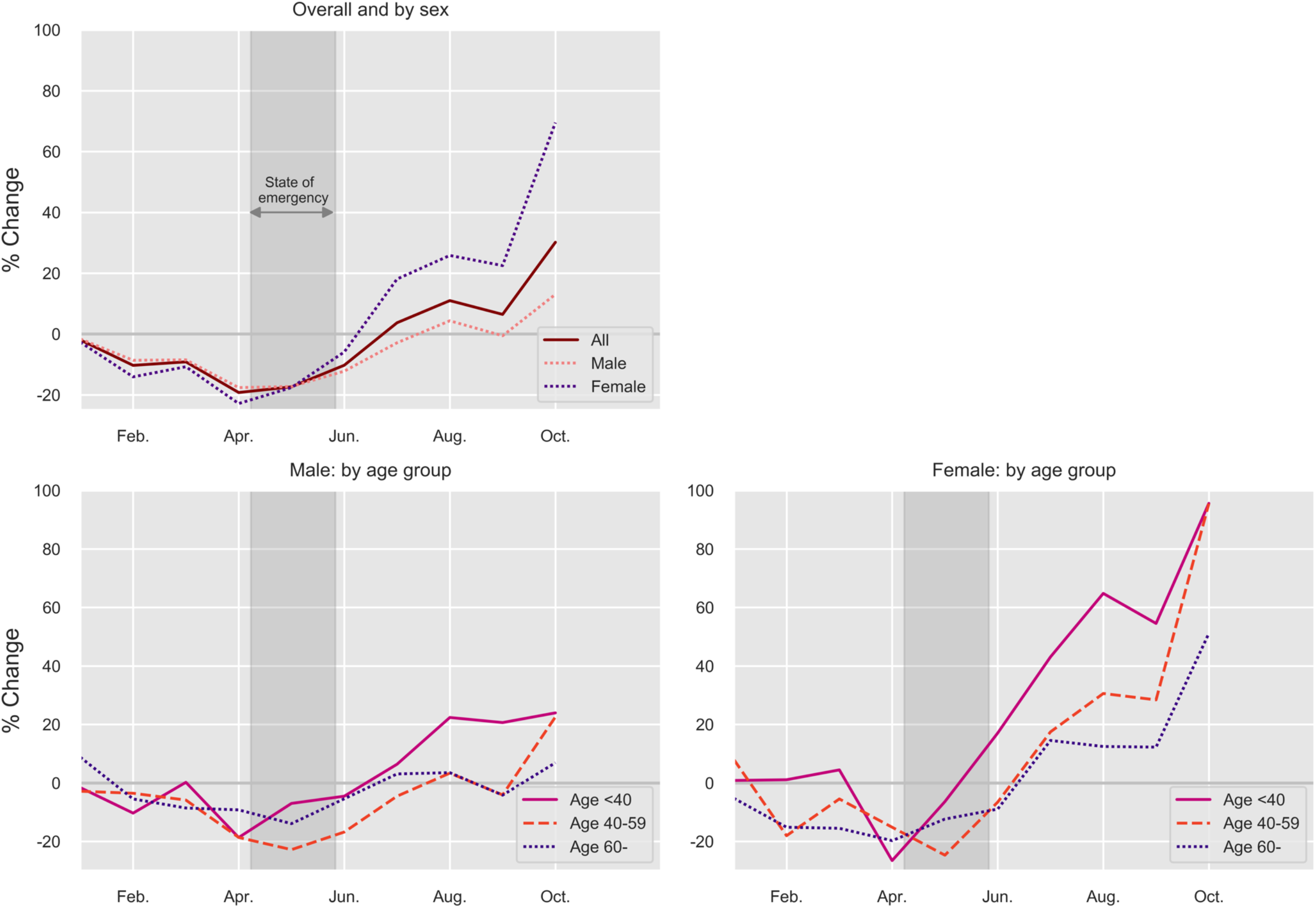
The percentage changes in suicide in Japan between 2020 and 2017-2019.

**Supplementary Fig. 4:**
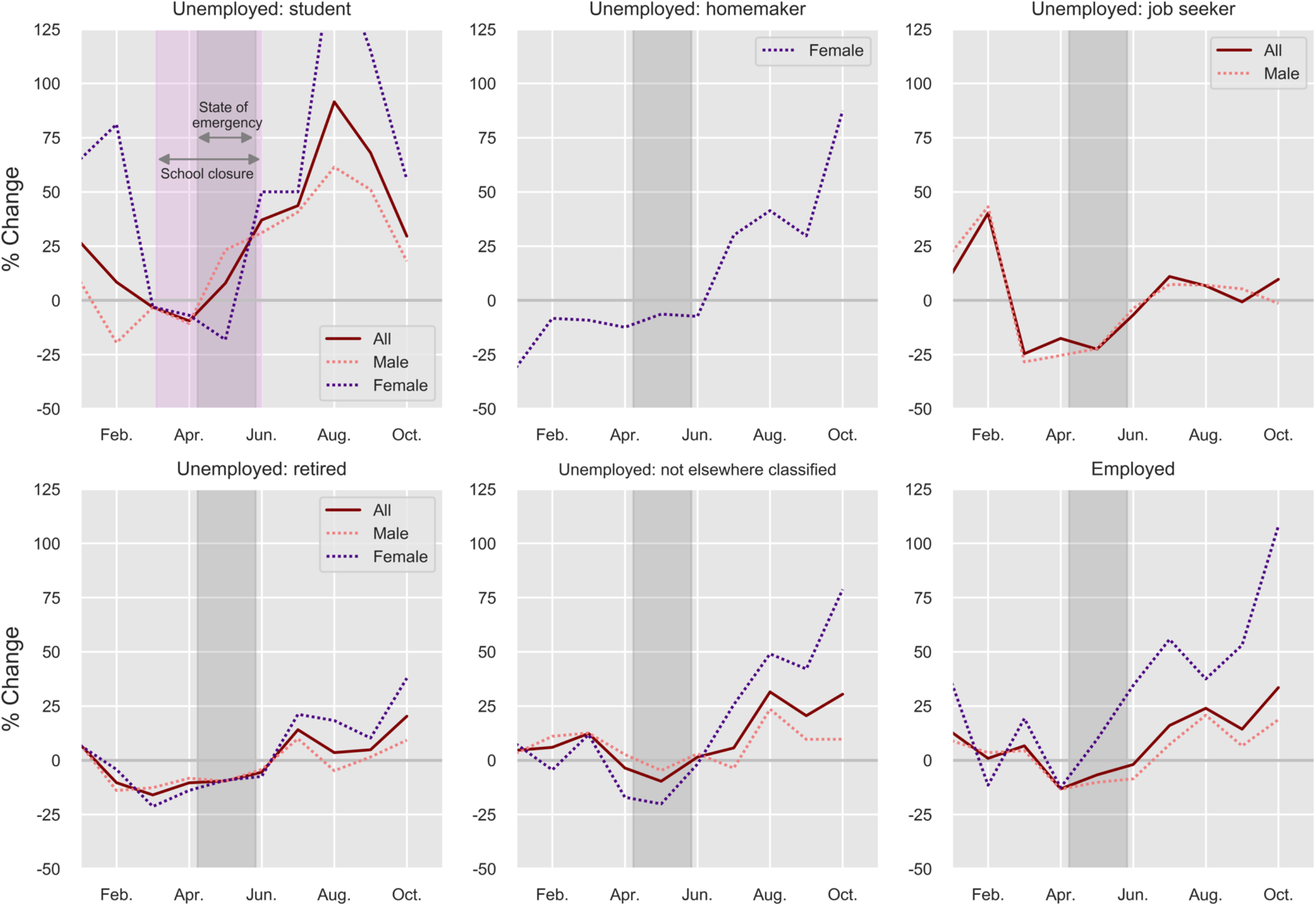
The percentage changes in suicide in Japan between 2020 and 2017-2019 by major occupation.

